# Estimating the maximum risk of measles outbreaks due to heterogeneous fall in immunization rates

**DOI:** 10.1101/2023.10.24.23297486

**Authors:** Nicholas Wu, Sifat Afroj Moon, Ami Falk, Achla Marathe, Anil Vullikanti

**Affiliations:** Department of Mathematics, University of Maryland; Biocomplexity Institute, University of Virginia; Department of Psychology, University of Virginia; Department of Public Health Sciences, University of Virginia; Department of Computer Science, University of Virginia

## Abstract

Immunization rates for childhoold vaccines, such as MMR, have seen a reduction over the recent years; this fall has only been accentuated after the COVID-19 pandemic. However, there is limited data on where the rates have reduced, and prior work has shown that heterogeneity in the drop in immunization rates has a significant impact on the risk of an outbreak. An important question from a public health perspective is: what is the maximum size of an outbreak in a region, when limited information is available on the fall in immunization rates within the region?

This turns out to be a very hard computational problem. We develop a Bayesian optimization based approach for estimating the maximum outbreak size, and use it on a measles model for the state of Virginia. Our results show that the maximum outbreak size is several orders of magnitude higher than estimated in a baseline which assumes homogeneous fall. Even for a 5% reduction in the statewide immunzation rate, the expected outbreak size can be very high. The maximum outbreak size depends crucially on the importation location, i.e., where the disease starts, and importation in an urban region leads to a significantly higher outbreak. The outbreak size remains high even if the drop in immunization is bounded in health service areas in the state.

## 1 Introduction

Measles is one of the most contagious childhood diseases, with a high reproductive rate (*R*_0_), often reported to be in the range 12 − 18 [21]. Fortunately, measles is a vaccine-preventable disease by the Measles, Mumps, and Rubella (MMR) vaccine. However a high level of immunization rate (more than 95%) is required to reach herd immunity. MMR is a required vaccine in the US, and in most parts of the world but compliance remains an issue due to multiple factors such as religious beliefs, access to health care, vaccination fees, misinformation about vaccine safety, lack of transportation etc.

In the US, MMR vaccination rates have been high enough for herd immunity and in year 2000, measles was declared as “eliminated” from the US [10]. “Elimination” means that the disease is no longer constantly present in the country but travelers can still bring the disease and these imported cases can cause an outbreak among the ones who are undervaccinated or unvaccinated. Additionally, there has been growing vaccine hesitancy in the US in the past two decades.

As a result, US has observed multiple measles outbreaks in the last few years. For instance, a large outbreak in New York (NY) in 2019 caused over 900 cases in NY, and over 1200 cases overall in the US [36]. The risk of measles, and other vaccine-preventable diseases, has significantly been exacerbated due to the COVID-19 pandemic [43]. In the US, as well as in many parts of the world, there has been a significant drop in routine immunizations during the pandemic, e.g., [38, 9, 24]. For instance, Seither et al. [38] noted that during the 2020–21 school year, national coverage of required vaccines (which includes MMR) among kindergarten students declined from 95% to approximately 94%. Over 27 million children were estimated to have missed the first dose of the measles vaccine in 2020 [9], and measles is now viewed as an imminent global threat [24]. Therefore, understanding the risk of a measles outbreak due to a drop in immunization rate is an important public health problem, and is the focus of our paper. We refer to this as the MaxRisk problem, and formalize it in Section 2.2.

There are two significant difficulties in addressing the MaxRisk problem. First, data on immunizations is available at limited level of detail. Surveys that provide estimates of drop in immunization rates have been reported done at spatially coarse levels, e.g., [38, 9], which was also noted in [28]. Immunization rates have been known to be quite heterogeneous, and underimmunized spatial clusters have been reported [35, 8, 27, 14, 1]. It is quite likely that the drop in immunization is also quite heterogeneous. Masters et al. [28] study this problem using a simple epidemic transmission model and show that coarse levels of spatial aggregation of non-vaccinated clusters can grossly underestimate the risk of outbreaks. Hence finer resolution data is needed to correctly identify target regions for intervention.

If immunization rates are known accurately at a high resolution, an epidemic transmission models can be used in getting bounds on the outbreak. However, if the immunization rates are only known at a coarse level (e.g., at the level of a state), the outbreak risk can be formalized as the maximum outbreak size over the space of all high resolution immunization rate vectors consistent with the state level, as initiated in [28]; however, they consider a simple epidemic model, for which the maximum is easy to solve. The second big challenge in the MaxRisk problem in realistic epidemic models (e.g., [45, 13, 18, 43], which account for the heterogeneity in the population and mixing) is that finding the maximum turns out to be a very challenging computational problem. The decision space is very large and high dimensional, and the objective function is non-convex and stochastic, making it very difficult to solve.

In this paper, motivated by the approach of [28], we formalize the MaxRisk problem to estimate the maximum expected outbreak size under a known decline in immunization rate. We develop a novel computational approach, LAMCTS-EPI, to bound the outbreak size due to a reduction in vaccination rates, using Bayesian optimization techniques. We apply our approach to obtain estimates for Virginia under varying levels of immunization drop. We find that the outbreak risk estimated by LAMCTS-EPI is significantly high, much higher than a baseline which assumes proportional drop in vaccination. We also observe that the importation location, i.e., where the outbreak starts, has a significant impact on the overall outbreak size, but the worst case solutions have complex structure, and do not have good correlation with population size and other county level features.

## 2 Methods

### 2.1 Disease transmission model

A number of mathematical models have been used for studying different kinds of questions related to measles, e.g., [17, 13, 45, 20]. Here, we use the gravity tSIR (Time-Series Susceptible-Infected-Recovered) model for modeling the spread of measles in Virginia; *we note that the framework developed in this paper is applicable to any mathematical disease model*. The tSIR model was introduced in [45], and has been used in a number of studies on measles, e.g., [13, 18]. It belongs to a class of models known as metapopulation models, because it models a “population of populations”. The region of interest, denoted by ℛ is divided into smaller subregions (corresponding to zip codes), ℛ = {*r*_1_, *r*_2_, …, *r*_*n*_}. Let 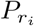 denote the population of subregion *r*_*i*_, and let dist(*r*_*i*_, *r*_*j*_) denote the distance between subregion *r*_*i*_ and subregion *r*_*j*_. The model assumes that each subpopulation 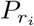 mixes homogeneously. Mixing across the patches (subregions) is modeled by the gravity model, which assumes that movement scales proportionally to a functional form analogous to Newton’s law of gravitation; this is discussed more below, and is fully described in Appendix **??**.

The tSIR model considers individuals to be in susceptible *S*, infected *I*, and recovered *R* states, and describes the evolution of the number of individuals in each state. The model is described completely in Appendix **??**. We assume zero mortality and complete immunity on recovery. Further, one timestep is assumed to correspond to the latent and infectious period of a case of measles, which is two weeks. Some portion of the population in each region is assumed to be immunized (which is needed as an input). We assume the vaccine has 100% efficacy in the model.

#### MMR vaccination coverage in Virginia

Immunization rates for all zip codes are needed as an input for the tSIR model. To find the vaccine coverage in Virginia, we use the publicly available Virginia School Immunization Survey (SIS) report for fall 2018 [30] and National Immunization Survey reports from the CDC’s ChildVaxView program. However, a challenge is that this dataset gives school level immunization, while our model needs zip code level rates. To get the zip code level vaccination rate from these two survey reports, we combine the SIS with a synthetic contact network, as described below.

We use a detailed activity based social contact network *G*(*V, E*) for Virginia developed by [3, 12, 16], where each node represents an individual, and each edge represents co-location based contacts. The network *G* has around 7.6 million nodes. Each node has rich demographic and geographic properties, such as age, gender, household size, and location. These attributes are statistically equivalent to the original census data when aggregated to a census block group level [2, 4]. This population contact network *G* is developed from various commercials and public data sources, including: 1) 2018 US census data, 2) American Time Use Survey data [42], 3) National Household Travel Survey Data [32], and 4) Multinational Time Use Study [31]. Each node or synthetic individual is part of a household, and has contacts with other synthetic individuals [2]. Further, each individual is assigned a daily set of activities (e.g., work, school, home, travel etc.), consistent with time use surveys.

To compute the vaccine rate at the zip code level, we assign vaccine status to nodes in the network *G*, then aggregate the network *G* to a zip code level. We calculate immunization status for children (up to age 17) from the publicly available Virginia School Immunization Survey (SIS) report for Fall 2018 [30]. The Virginia Department of Health (VDH) publishes the SIS report, which contains MMR immunization records for all public schools except kindergarten-level schools and schools with less than ten students. The immunization rate among children younger than twelve is assigned from the associated school’s kindergarten immunization rate. If an MMR-specific rate is unavailable for a school, we use the overall immunization rate.

Each individual in *G* is assigned an immunization probability in the following manner

- Each school age individual in the network model *G* is mapped to a school (unless they are homeschooled); school information is used from the National Center for Education Statistics database. We match each school corresponding to an individual in *G* by name to a school in the SIS data, which allows us to assign SIS immunization rates to specific schools; this is used to assign an immunization probability for each individual in *G* associated with that school. If a school in network *G* is not present in the SIS dataset, we use the available MMR immunization rate from the nearest neighboring school (using its latitude and longitude).
- The vaccine status among adults in the network *G* comes from the National Immunization Survey (NIS) reports from the CDC’s ChildVaxView program. It contains immunization rates among 19-35 months old from 1995 to 2017 at the state-level [23]. We estimate the immunization rate for adults using NIS data from 1995-2004 since adults were children in that time frame. Finally, we aggregate *G* at the zip code level to get the vaccine rate.

#### Model calibration

This includes two parts: the gravity model, and the disease transmission model. We first calibrate the gravity model, and then using the fitted gravity parameters, we calibrate the transmission parameter of the disease model, as described below.

##### Calibrating gravity model

The 2011-2015 5-year American Community Survey commuting flow data was used to calibrate the gravity model [7]. These data tables provide estimates for the number of workers who commute from one county to another within the United States. However, in this study, the gravity tSIR model is set up to model zipcode-level human movement. To calibrate our zipcode-level model using county-level commuter data, we obtain county-level predictions using gravity model by aggregating human movement flows up to the county level from the zip code level. This is achieved by geocoding each zip code using OpenStreet Nominatim [34], and extracting the county it belongs to from its address. Then, we obtain county-level flows by summing all flows from zip codes in the originating county to zip codes in the destination county. To obtain the final set of gravity model parameters, we then minimize the mean squared error between the gravity model county-level predictions and the county-level commuter flow data using L-BFGS as implemented in scipy; we note that due to technical reasons, the specific model we use is slightly different from that of [45]. A more detailed description of the procedure, along with supporting figures, is in Appendix **??**.

##### Calibration of the disease model

After calibrating the gravity model, we calibrate the transmissibility parameter of the tSIR model, *β*, which roughly corresponds to *R*_0_. Since there is no recent measles outbreak data for the state of Virginia, we calibrate against data from the 2018-2019 New York City measles outbreak by varying the parameter *β* until the resultant outbreak is roughly proportional to that of the NYC outbreak. More details can be found in Appendix **??**

### 2.2 Problem statement

The key focus of our paper is the following question: *if the statewide vaccination coverage is known to have declined by a certain percentage, what is the maximum risk of a measles outbreak that is possible?* Given that many measles outbreaks can die out quickly or are extremely small, we will consider two metrics for quantifying outbreak risk: the expected size of an outbreak and the probability of a large outbreak (when it exceeds a given threshold value). Solving these optimization problems involves considering the space of all possible immunization declines across all zip codes (while satisfying the total decline at the State level), which is a very challenging combinatorial optimization problem. In order to understand the importance of surveillance, we also consider the setting in which the decline is constrained within each region; for concreteness, we consider HSA (Health Service Areas) regions in Virginia. We describe the problems formally below.

#### 2.2.1 The MaxRisk problem

Informally, the MaxRisk problem involves determining the maximum expected outbreak size (in the tSIR model) if the total decline in immunization rate in the entire region is *α*%. We assume the expected outbreak size is estimated by the tSIR model, once the decline in each region *r*_*i*_ (a zip code) is specified, subject to the constraint that the total decline is *α*%. Therefore, the MaxRisk problem involves exploring the space of vectors which specify the decline in each region, with this constraint. This optimization problem turns out to be very challenging, therefore, we formalize it as a two level problem: (1) we consider a partitioning of the region into counties, and of each county into zip codes, and assume the decline within all zip codes contained in a county is constant, (2) we coarsen the decision space to specify the declines for each county. Note that the hierarchical partitioning into counties and zip codes can easily be changed to some other units. Also, the problem can be solved at the zip code level itself, but it is computationally much harder.

Let *Z* = {*z*_1_, …, *z*_*n*_} and *C* = {*c*_1_, …, *c*_*m*_} denote the set of zip codes and counties within the entire region of interest (the state of Virginia, in our study). Each zip code is part of a county, specified by the mapping *M* : *Z* → *C*, so that *M*(*z*_*i*_) = *c*_*j*_ if zip code *z*_*i*_ is in county *c*_*j*_. Let *V* and *P* be the vectors encoding the respective county-level initial vaccination rates and population; i.e 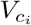 and 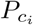 denote the overall vaccination rate and population of county *c*_*i*_, respectively. Analogously, let *V* ^*Z*^ and *P*^*Z*^ be the vectors encoding the zip code-level vaccination rates and populations, respectively.

To formalize the problem, we define a decision vector *x* ∈ [0, 1]^*m*^, which specifies the county-level vaccination decline, i.e., 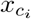 is the percent decline in county *c*_*i*_. As mentioned earlier, for computational feasibility, we assume the decline in each zip code *z*_*j*_ within county *c*_*i*_ is equal to 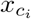. A decision vector *x* is feasible if it satisfies the following two conditions:

- The total decline across the state is at most *α*%, where *α* is a parameter. Formally, 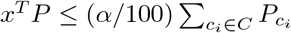
- For each county *c*_*i*_, the decline in each zip code is non-negative, i.e., 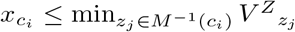, where *M* ^−1^(*c*_*i*_) is the set of all zip codes belonging to county *c*_*i*_.

Our objective is to find *x*, which results in the largest expected outbreak size. Let 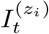 denote the number of infections in zip code *z*_*i*_ at time *t*. Then, our objective is the expectation of 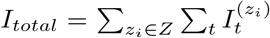; we approximate it using *N* simulation runs of the tSIR model. The number of infections is a function of the disease model parameters, denoted by *θ* (which also specifies the importation), the current immunization rate *V*, and the declines *x* (note that *V* − *x* specifies the county level immunization rate); we use *f*(*x, V, θ*) to denote 𝔼[*I*_*total*_], the expected number of infections. Putting everything together, the MaxRisk problem can be formally stated as

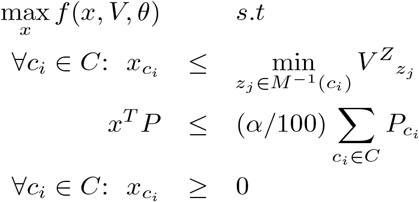

We note that the problem can be defined at the zip code level by considering *Z* = *C* in the above formulation. Another important parameter in the tSIR model is the seeding of the disease. Our results show that the outcomes are highly sensitive to seeding strategies.

#### 2.2.2 MaxRiskConstrained problem

We now consider an extension of the problem, where information on declines is available for different parts of the state (e.g., through surveys), referred to as *surveillance regions* 𝒮 = {*s*_1_, …, *s*_ℓ_ }. We assume is a coarser partitioning of the region than *C* (the set of counties), so that each county is contained in some surveillance region. Let *M*_*s*_ : *C* → 𝒮 be the mapping of counties to surveillance regions, so *M*_*s*_(*c*_*j*_) = *s*_*i*_ implies that *c*_*j*_ is a part of *s*_*i*_. Let *α*_*i*_ denote the reported percentage decline in immunization rate in the region *s*_*i*_. The MaxRiskConstrained problem involves optimizing over the space of vaccine decline vectors *x*, which satisfy the constraint that the decline within *s*_*i*_ is at most *α*_*i*_%, and the total state level decline is *α*%. This is formalized below.

We continue with the two level regions *Z* and *C* (interpreted as zip codes and counties), defined in Section 2.2.1 in the case of the MaxRisk problem. We now say that a decision vector *x* is feasible if it satisfies the following two conditions:

- For each surveillance region *s*_*i*_, the total decline across *s*_*i*_ is at most *α*_*i*_%. Formally,

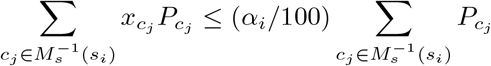
- For each county *c*_*j*_, the decline in each zip code is non-negative, i.e., 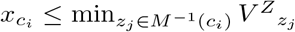, where *M*^−1^(*c*_*i*_) is the set of all zip codes belonging to county *c*_*i*_.

As before, let *f*(*x, V, θ*) denote the expected outbreak size, where *x, V* and *θ* are the decline vector, immunization rate, and disease model, respectively. The MaxRiskConstrained can now be formally defined as

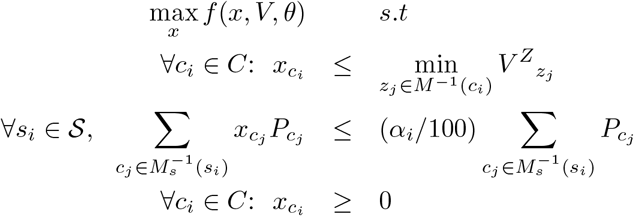

In our simulations, we consider HSA regions in Virginia to be the surveillance regions; our methods can be extended to any other partition just as easily.

For convenience, we will use the notation 𝒳 to refer to the feasible sets of both MaxRisk and MaxRiskConstrained. The most important structure of the feasible sets of MaxRisk and MaxRiskConstrained are that they are polytopes (i.e defined solely by linear constraints). Since our solver (described in the next section) can handle any problem defined over a polytope, we will use 𝒳 to interchangeably refer to the feasible sets of both MaxRisk and MaxRiskConstrained.

### 2.3 Our algorithm for solving MaxRisk: LAMCTS-EPI

The MaxRisk and MaxRiskConstrained problems formulated above have a number of features that make it challenging to solve using standard off-the-shelf solvers. Namely, (1) they are relatively high-dimensional (i.e. hundreds of dimensions); (2) the objective function is expensive enough that many repeated evaluations would be costly; (3) gradient information is not readily available, and the problem is non-convex.

Standard Bayesian optimization (BO) can fulfill criteria (2) and (3) above. It can be used to solve problems of the following form: max_*x*∈Ω_ *f*(*x*) where *f* is a function that is expensive to evaluate, does not have a closed form, and has no gradient information; and Ω is a set with some known structure, e.g a rectangular set Ω = {*x* ∈ ℝ^*d*^ | *l*_*i*_ ≤ *x*_*i*_ ≤ *u*_*i*_} for some bounds 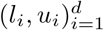. For more background, see the tutorial by Frazier [19].

The basic idea of Bayesian optimization is to construct a statistical model 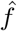 of the unknown function and use the knowledge from the model to select new points to acquire in a resource-efficient manner. The model is usually obtained via Gaussian Process (GP) regression, which is a Bayesian model that provides a posterior distribution over probable functions. Practically speaking, we can think of the model as providing uncertain estimates for each *x* ∈ Ω in the form 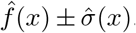. To perform optimization, we begin by sampling the domain Ω and constructing a dataset of function evaluations 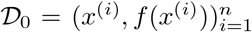. We use 𝒟_0_ to fit an initial model 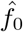. Then, the next point to evaluate *x*^(*n*+1)^ is selected by an *acquisition function*, which is a rule that helps balance between exploitation of current knowledge about the function and exploration of possibly unobserved optima. Once a new pair (*x*^(*n*+1)^, *f*(*x*^(*n*+1)^)) is evaluated, we augment our dataset: 𝒟_1_ = 𝒟_0_ ∪ {(*x*^(*n*+1)^, *f*(*x*^(*n*+1)^)}. The augmented dataset is used to fit a new model 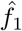, which we can use to re-compute the acquisition function and select a new point. The process repeats until a desired number of samples are collected or until the computational budget is exceeded.

Common acquisition functions include Upper Confidence Bound (UCB), expected improvement (EI), and Thompson sampling (TS). The idea of UCB is to select the next point as the point with the largest potential upper bound, i.e 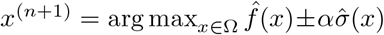, where *α* is a tunable paramter. EI chooses the next point so that the expected gain over the current optimum is maximized. If the current optimal objective value is 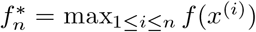, then we choose the next point as 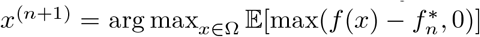 (where the expectation is taken with respect to the posterior distribution induced by 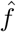.) Thompson sampling is a stochastic acquisition function, which works by sampling a function *g* from the posterior distribution induced by 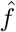. The next point is then *x*^(*n*+1)^ = arg max_*x*_ *g*(*x*). In practice, since the function *g* is infinite-dimensional, TS for BO is approximated by sampling the function *g* at a finite number of points and taking the max of the samples. Expected improvement is discussed by Frazier [19]. Shahriari et al. discuss Thompson sampling in the bandit case (finite domain) [39]. Practical assessment of EI, UCB and other acquisition functions is given in Snoek et al. [41]

However, standard BO is difficult to apply here given criteria (1), as performance is known to suffer in higher dimensions. For instance, a known problem is over-exploration, especially at boundaries of the search domain [33]. Over-exploration wastes the computational budget by searching excessively in regions that have already been sampled. Thus, alternate approaches are needed to apply BO to the problem at hand.

In this study, we modified a Bayesian optimization framework known as Latent Action Monte Carlo Tree Search (LAMCTS) [44], which is a meta-algorithm that helps BO search within high-dimensional spaces. We call the new algorithm LAMCTS-EPI, which we use to solve MaxRisk and MaxRiskConstrained. In these problems, recall the feasible set 𝒳 is the set of all feasible *x* vectors, and the objective *f*(*x*) = *f*(*x, V, θ*) is the expected outbreak size. Notice that for both problems MaxRisk and MaxRiskConstrained, the set 𝒳 is a polytope, i.e defined by linear constraints. Thus, we tailor our method for problems with linear constraints.

The idea of LAMCTS-EPI is to control exploration of the feasible scenarios 𝒳 by balancing global search and local exploration. Global search is controlled by recursively partitioning 𝒳 into “good sets” and “bad sets” during optimization. “Good sets” are expected to contain *x* such that *f*(*x*) is high (large outbreaks), and “bad sets” are expected to contain *x* such that *f*(*x*) is low (small outbreaks). The division of the region is described by a hyperplane which bifurcates the feasible region. This recursive partitioning, represented by a search tree, can be explored by Monte Carlo Tree Search (MCTS) for further local exploration.

For this local exploration step, LAMCTS-EPI uses a variant of BO called TurBO (Trust Region Bayesian Optimization) [15], which avoids over-exploration by constraining search to a local trust-region. To focus on local exploration, TurBO maintains a bounding box (the trust region) centered at the best point observed so far. It then maintains a model within the bounding box, and searches for promising points to acquire. If several new points that are evaluated within the box beat out the current optimum, TurBO expands the trust region to try to explore more. If TurBO fails to find new points that beat the current optimum, then TurBO shrinks the trust region to focus on searching around the current optimum. TurBO terminates when the trust region shrinks beyond a predefined threshold. More details can be found in the original LAMCTS paper by Wang et al. [44], and the TurBO paper by Eriksson et al. [15].

Our contributions in LAMCTS-EPI are as follows:

- *Linear constraints*. The original description of LAMCTS by Wang et al. only considered the case of rectangular domains. We expand LAMCTS-EPI to allow the case where the feasible set is a polytope, e.g {*x* ∈ ℝ^*d*^ |*Ax* ≤ *b* and *l* ≤ *x* ≤ *u*} for some matrix *A* ∈ ℝ^*m×d*^, for some vector *b* ∈ ℝ^*m*^, and for a pair of upper and lower bound vectors *l, u* ∈ ℝ^*d*^. Such constraints specially arise in the MaxRiskConstrained problem, where we have a constraint for each surveillance region. Handing
- these constraints is done by restricting sampling of initial points and subsequent candidate points to the feasible set; described further in the next bullet point.
- *Hit-and-run sampler*. LAMCTS uses accept-reject sampling to produce candidate points to sample partitions. Accept-reject is a general sampling procedure that can be used to draw samples in regions defined by non-linear boundaries. However, it is prohibitively expensive to use in high-dimensional spaces due to the curse of dimensionality. In fact, in our initial tests of LAMCTS with accept-reject sampling, a personal computer could not complete one iteration of sampling within several hours on a problem of modest dimension (e.g 10 dimensions). On the other hand, if one enforces a partitioning of the feasible set 𝒳 by hyperplanes rather than arbitrary nonlinear boundaries, all the constraints that define candidate regions (including those from the problem itself) become polytopes rather than of arbitrary shape. This makes sampling tractable by enabling the usage of fast algorithms for sampling polytopes, in particular the *hit-and-run algorithm*. The hit-and-run algorithm is a Markov Chain Monte Carlo (MCMC) based algorithm which simulates a random walk on the polytope [40]. The random walk eventually converges to the uniform distribution on the polytope [5]. We utilize the implementation of the “hit and run” algorithm in `hopsy` [37], a Python wrapper for the sampling package HOPS [25] to sample regions created by LAMCTS-EPI. With `hopsy`, sampling the aforementioned feasible region now completes in a few seconds on a personal computer.
- *Parallelization and batch acquisition*. The original TurBO paper by Eriksson et al. [15] demonstrated that using batched Thompson sampling [22] as the acquisition function can result in essentially linear speedups in computation time. In batched Thompson sampling, instead of sampling and optimizing one model *g* from the posterior distribution induced by 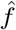, we sample *q* models *g*_1_, …, *g*_*q*_ and evaluate *f* at each of the points *x*^(*n*+1)^, …, *x*^(*n*+*q*)^ from the models. Thus, we parallelized the tSIR simulation and used batched TS as the acquisition function for LAMCTS-EPI.

A basic workflow of LAMCTS-EPI is given in Figure 1. Our objective is to maximize the expected outbreak size, *f* : 𝒳 → ℝ, where 𝒱 ⊆ [0, 1]^*d*^. The feasible set of MaxRisk/MaxRiskConstrained, 𝒳, is a polytope, and so we write 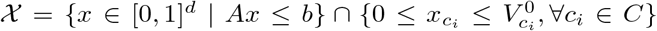 for a given matrix *A* ∈ ℝ^*m*×*d*^ and *b* ∈ ℝ^*m*^, and the initial vaccination vector *V* ^0^.

**Figure 1:**
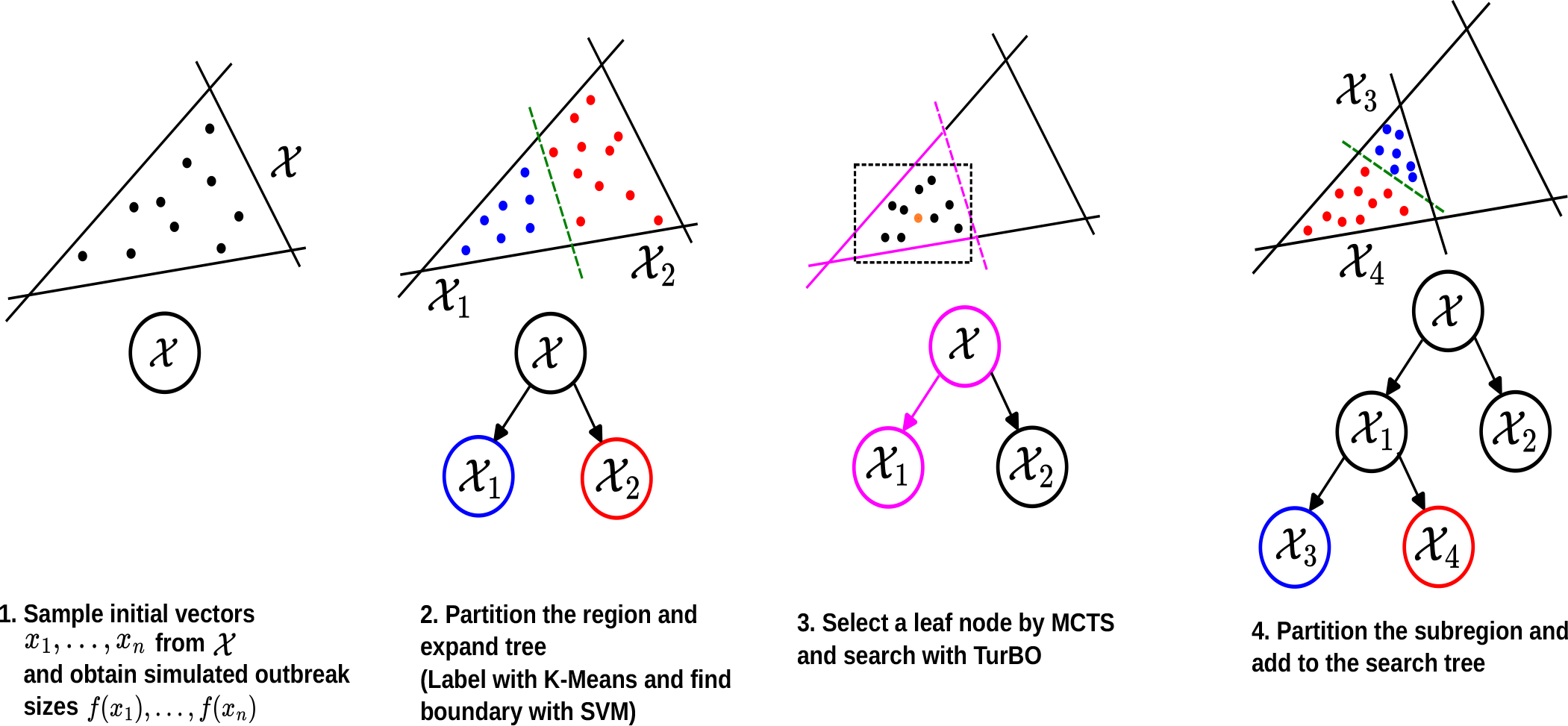
Schematic showing a simplified example iteration of LAMCTS-EPI. In MaxRisk and MaxRiskConstrained, the feasible set (possible *x* vectors) is Ω =𝒳, which is a polytope. The objective is to maximize the expected outbreak size as a function of *x*, which we denote by *f*(*x*) (dropping the immunization rate vector and the disease model parameters, for convenience). To optimize, first we sample and evaluate candidate vectors *x*^(1)^, …, *x*^(*n*)^ using a hit-and-run algorithm (`hopsy`) over 𝒳. We compute *f*(*x*^(*i*)^) for 1 ≤ *i* ≤ *n*, and then divide 𝒳 into a “good” or “large outbreak” (blue) region; and a “bad” or “small outbreak” (red) region. This division grows the search tree. Then, we perform Monte Carlo Tree Search (MCTS) on the search tree to select a new subset to search locally within. LAMCTS-EPI traverses the tree from the root, selecting children according to their potential to hold “good” points until it reaches a leaf node, quantified by a UCT (Upper Confidence bound for Trees) rule. Once a subset (leaf node) is selected, local search is carried out by TuRBO (Trust Region Bayesian Optimization), which maintains a shrinking bounding box around the best candidate solution. We again use `hopsy` to draw candidate points from the intersection of the chosen subset and the trust region. After TurBO finishes, the evaluated points are used to split the region further, and the algorithm repeats the process of searching and expanding the tree.

A typical run of LAMCTS-EPI begins by instantiating the global search tree. First, we sample *n* decision vectors *x*^(1)^, …, *x*^(*n*)^ from the uniform distribution over 𝒳 using `hopsy`, which takes as arguments the constraints defined by matrix *A* and vector *b*. We run the simulation for each sampled *x*^(*i*)^ and obtain outbreak sizes, constructing the initial dataset 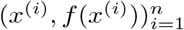.

The global search tree is constructed by partitioning this dataset. To partition, we build feature vectors by concatenating the outbreak size to the *x* vectors, yielding [*x*^(*i*)^, *f*(*x*^(*i*)^)] ∈ R^*d*+1^ and running K-means with *k* = 2 on this dataset. We then fit a linear SVM to separate the data according to the labels from K-means. The total number of partitioning steps carried out (and hence the depth and size of the resulting tree) is controlled by the *leaf size*, a tunable hyperparameter which designates the maximum number of samples that can be collected in a subregion before it becomes eligible to be split. As the feasible set 𝒳 is split, one region is designated as the “good” region, e.g 𝒳 _1_, and the other the “bad region,” e.g 𝒳 _2_. To determine the “good” region, we first compute the mean outbreak size of the points in a region. Let 𝒟(𝒳_*j*_) denote the set of collected evaluations (*x*^(*i*)^, *f*(*x*^(*i*)^)) such that *x*^(*i*)^ ∈_*j*_ 𝒳_*j*_. Then Mean 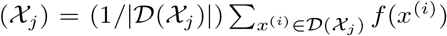. Whichever region has the higher mean outbreak size is designated the “good” region.

Then, the global search is carried out by selecting a leaf node (candidate region) of the tree by MCTS. This is performed by starting at the root. We select one of the two child nodes as the next node, and repeat the process of choosing child nodes until we reach a leaf node. The child nodes are selected by a UCT (Upper Confidence bound for Trees) rule. The UCT rule performs a similar role to the acquisition function in BO, balancing exploitation of known high-performing regions with exploration of underexplored regions that may possibly contain larger-outbreak-size. To compute the UCT score of region *𝒳*_*j*_, suppose that its parent node is 𝒳_*p*_ and let *N* (𝒳_*j*_) denote the number of times region *X*_*j*_ was visited in the tree search. Then, we have 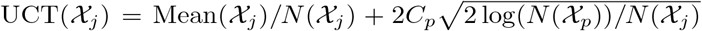 where *C*_*p*_ is the *exploration factor*, a tunable parameter. A higher *C*_*p*_ encourages more exploration. MCTS selects the child node with the highest UCT value.

Once MCTS selects a leaf node, we perform local search of the selected subregion with TurBO. TurBO initializes a local model over the region defined by the leaf node, and begins acquisition of candidate points within the trust region via batch Thompson sampling. We use `hopsy` again to sample candidate vectors from the intersection of the trust region and the selected node. As TurBO continues to search within the region, it collects more sample pairs (*x*^(*i*)^, *f*(*x*^(*i*)^)). After TurBO terminates, these samples are added to the dataset, and the tree is updated. More regions can be created by splitting, dependent again on the splitting threshold and the region which TuRBO samples in. This deepens the search tree, and then the algorithm repeats, choosing a leaf node to search once more by MCTS. A complete description of LAMCTS-EPI and the modifications are described in Appendix **??**.

### 2.4 Specific questions and experimental setup

We aim to understand the following questions:

1. How much worse are the LAMCTS-EPI scenarios relative to a natural baseline scenario, which has a constant decline of *α* in all counties?
2. What is the effect of *α* and the importation region on the the outbreak size?
3. What is the difference between MaxRisk and MaxRiskConstrained, i.e., if more surveillance information is available, how is the outbreak risk affected?
4. Is there any structure or pattern (e.g spatial) in the solutions produced by the LAMCTS-EPI?

#### The Proportional heuristic

For a given *α*% decline, we assume that the decline in each zip code is *α*% decline. In the notation of MaxRisk, we would write this solution as *x*_Proportional_ = [*α, α*, …, *α*]. We can then estimate the severity of outbreaks from LAMCTS-EPI obtained vectors *x*_LAMCTS-EPI_ by comparing the expected outbreak sizes *f*(*x*_Proportional_) and *f*(*x*_LAMCTS-EPI_) for a fixed initial condition and *α*.

#### Scenarios and parameters in the experimental setup

We consider the following four classes of scenarios. The first three are for the MaxRisk problem, and are defined in terms of case-importation; the decline in immunization is assumed to be *α*% in the entire state, where *α* is a parameter. For each of these, we have a scenario corresponding to declines based on the Proportional and LAMCTS-EPI solutions. The fourth is for the MaxRiskConstrained problem.

1. **Dulles Proportional and Dulles LAMCTS-EPI**: One initial case is imported in the Virginia zip code 20166, where the Dulles International Airport (IAD) is located.
2. **Highland Proportional and Highland LAMCTS-EPI**: One initial case begins at Virginia zip code 24465 in Highland County, which has the lowest population density in Virginia.
3. **Random Proportional and Random LAMCTS-EPI**: One initial case is placed in one randomly selected zipcode. The zipcode is selected uniformly and randomly from the dataset of 705 Virginia zipcodes. This process is repeated 25 times, resulting in 25 distinct seeding locations. We analyze the Proportional and LAMCTS-EPI scenario for each of these seeding locations.
4. **HSA**: This scenario corresponds to MaxRiskConstrained. The initial case begins in zip code 20166. Additionally, we assume *α*% decline in each of the 5 HSA regions in Virginia^1^.

The Dulles and Highland scenarios are intended to explore the impact of seeding in regions of varying population density, with the Northern VA region where Dulles Airport is situated, being one of the most dense and Highland County being the least dense in VA. The Random scenario is intended to understand the “average” scenario. The HSA scenario explores the extent to which improved surveillance can constrain the worst possible scenario.

Table 1 provides a summary of all different scenarios and parameters. For each scenario above, we vary the parameter *α* from 5 to 10 in increments of 1 for a total of 5 optimization runs per scenario. The disease model parameters were chosen using the calibration method described earlier.

**Table 1:** Summary of changing parameters for each scenario. Each row represents one concrete scenario. “Problem/heuristic” is the name of the rule or problem that determines how to find a solution for this setup. “Allocation strategy” provides a brief description of the solution strategy. “Seeding location” is the location of the initial case in all simulation runs under this scenario. ‘*α*’ denotes the range of the *α* parameter, the % allowable statewide vaccination decrease. ‘Total # of runs’ denotes how many total simulation or optimization runs were carried out for that scenario.

| Scenario | Problem/<br>heuristic | Allocation<br>strategy | Seeding<br>location(s) | $\alpha$ | Total # of<br>runs |
| --- | --- | --- | --- | --- | --- |
| Dulles Prop. | PROPORTIONAL | $\alpha\%$ decline<br>in all zip codes | zip code<br>20166 | 5, 6, ..., 10 | 5 |
| Dulles<br>LAMCTS-<br>EPI | MAXRISK | Chosen by<br>LAMCTS-<br>EPI | zip code<br>20166 | 5, 6, ..., 10 | 5 |
| Highland<br>Prop. | PROPORTIONAL | $\alpha\%$ decline<br>in all zip codes | zip code<br>24465 | 5, 6, ..., 10 | 5 |
| Highland<br>LAMCTS-<br>EPI | MAXRISK | Chosen by<br>LAMCTS-<br>EPI | zip code<br>24465 | 5, 6, ..., 10 | 5 |
| Random<br>Prop. | PROPORTIONAL | $\alpha\%$ decline in<br>all zip codes | 25 pre-<br>selected<br>random zips | 5, 6, ..., 10 | 125 |
| Random<br>LAMCTS-<br>EPI. | MAXRISK | Chosen by<br>LAMCTS-<br>EPI | 25 pre-<br>selected<br>random zips | 5, 6, ..., 10 | 125 (25 seeds,<br>5 $\alpha$ per seed) |
| HSA | MAXRISK<br>CONSTRAINED,<br>with $\alpha_i = \alpha$<br>for $i = 1, \dots, 5$ | Chosen by<br>LAMCTS-<br>EPI | zip code<br>20166 | 5, 6, ..., 10 | 25 (5 replicates<br>per $\alpha$) |

#### Summarizing outbreak distribution

To summarize the effects of the parameters on the characteristics of the outbreak distribution, we compute the following four estimates for all scenarios in Table 1. After obtaining the final vectors *x*_LAMCTS-EPI_ and *x*_Proportional_, we obtain a more refined estimate of the outbreak distribution by re-running the simulation parameterized by both *x* vectors and drawing outbreak samples *o*_1_, …, *o*_*N*_ with *N* = 10, 000 for all scenarios. The following are computed from these samples:

1. *Expected outbreak size*. As noted in Section 2.2.1, the expected outbreak size is the metric of interest for our optimization problem. Given the samples above, we compute it as 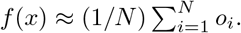. This is computed for both *x*_Proportional_ and *x*_LAMCTS-EPI_.
2. *Multipliers*. We use this to to quantify the gap between the Proportional solution and the LAMCTSEPI solutions. For *x*_Proportional_ and *x*_LAMCTS-EPI_ computed from the same initial conditions (or scenario; discussed in the previous subsection), the multiplier for that scenario is 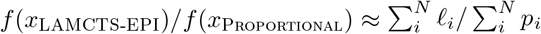, where ℓ_1_, …, ℓ_*N*_ are the LAMCTS-EPI samples and *p*_1_, …, *p*_*N*_ are the Proportional samples.
3. *Probability of outbreak*. We estimate the probability of an outbreak by computing the proportion of simulations where the outbreak size is above a selected threshold. If *t* is the threshold outbreak size, then we compute this quantity as 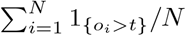; where 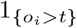 is the indicator function, returning 1 if the sample *o*_*i*_ exceeds the threshold *t* and 0 otherwise.

We compute this quantity for a lower *t* and a higher *t* to differentiate between the probability of outbreaks and the probability of *large* outbreaks.

- *Density estimates*. We compute density estimates of the outbreak distribution to understand qualitative aspects of the distribution that are not captured by the above statistics.

#### Clustering measures

We investigate the structure of obtained solutions by clustering measures and regression analysis. We use two metrics, Isolation Index and Moran’s-I [28] to understand the distribution of the county level declines in the LAMCTS-EPI solutions.

- Isolation Index: It measures the level of segregation within a specific group or cluster compared to the larger population. Formally, this is defined as 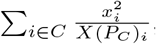, where *x*_*i*_ denotes the number of unimmunized people in county *i*, (*P*_*C*_)_*i*_ is the population of county *i*, as defined earlier, and *X* is the total number of unvaccinated people [28]. It can take values from 0 (no segregation) to 1 (full segregation).
- Moran’s-I: This is a measure of global spatial autocorrelation, which indicates how similar or dissimilar values are distributed across a spatial dataset. Formally, this is defined as 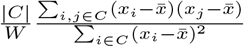, where *x*_*i*_ is the number of unimmunized individuals in county *i*, 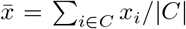 is its average, *w*_*ij*_ is the weight between counties *i* and *j*, and *W* =_*i,j*∈*C*_ *w*_*ij*_ [28]. Weight between two counties *i* and *j* is *x*_*i*_ is 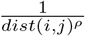. The value of Moran’s-I ranges from -1 to 1. A value of 0 indicates no spatial autocorrelation (or random distribution), while -1 represents clustering with dissimilar values (e.g., high and low values are clustered together), and 1 indicates clustering with similar values (e.g., high values are clustered with other high values).

#### Computational setup, procedure, and data

To perform optimization runs, we initialize the simulation model with the *β* parameter and gravity model parameters respectively calibrated from the NYC outbreak data and ACS flow data. Then, we begin the simulation at the seed specified by the scenario. Each time the simulation is run, it runs for 20 time steps, which corresponds to 40 weeks.

The objective function used in optimization for all scenarios is the expected outbreak size, which is approximated by the sample average outbreak size. We draw different *N* for optimization and for post-optimization data analysis. During optimization, for Dulles, Highland, and HSA; we draw *N* = 2000 simulations. The Dulles, Highland, and HSA optimization routines were run until the program terminated due to timeout after 168 hours or insufficient resources. Since there are many Random seeds, we draw a reduced number of samples, setting *N* = 500 during optimization. When optimization terminated, the best solution after exactly 1500 objective function evaluations was chosen as the representative solution. If runs terminated before reaching 1500 samples, the program was restarted and run until a total of 1500 samples were attained.

Since the HSA runs are compared to the Dulles results, we produced five runs for each *α* and took the best run out of the five as the final run for that *α* value, resulting in a total of 25 HSA runs. This is intended to avoid the possibility of artificially low HSA values due to local optima. We use the best out of the five runs for the final analysis.

For all scenarios, we evaluated final *x* vectors by re-simulating with *N* = 10000 simulation draws. This simulation dataset was used for estimates regarding the outbreak distribution, and we computed the mean outbreak size, the multipliers, the probability of an outbreak, and density estimates.

A complete description of settings is given in Appendix **??**.

## 3 Results

We discuss two classes of results. First, we study the impact of *α*, importation location, and different problem scenarios on different epidemic outcomes in Section 3.1. Next, we explore the geospatial structure of the critical regions, including the spatial statistics (Moran’s I and isolation index) in Section 3.2.

### 3.1 Effect on outbreak size and distribution

To understand the quality of the outbreak distribution, we plot the mean outbreak size, density estimates via violin plots, and the probability of outbreak (defined as the probability that the outbreak exceeds a fixed threshold). Figure 2 shows the mean outbreak size on a log scale for all the scenarios (optimizer and baseline) against *α*. Figure 3 shows the multipliers on a log scale for all scenarios against *α*. Figure 4 shows the outbreak distributions for each HSA region for *α* = 5, and also shows the maximum expected regional outbreak size for Dulles LAMCTS-EPI and HSA versus *α*. Figure 5 shows violin plots (density estimates) for all scenarios, with *α* on the x-axis. Figure 6 shows the outbreak probability for two different outbreak thresholds, 500 and 5000. We discuss the specific results from these plots below.

**Figure 2:**
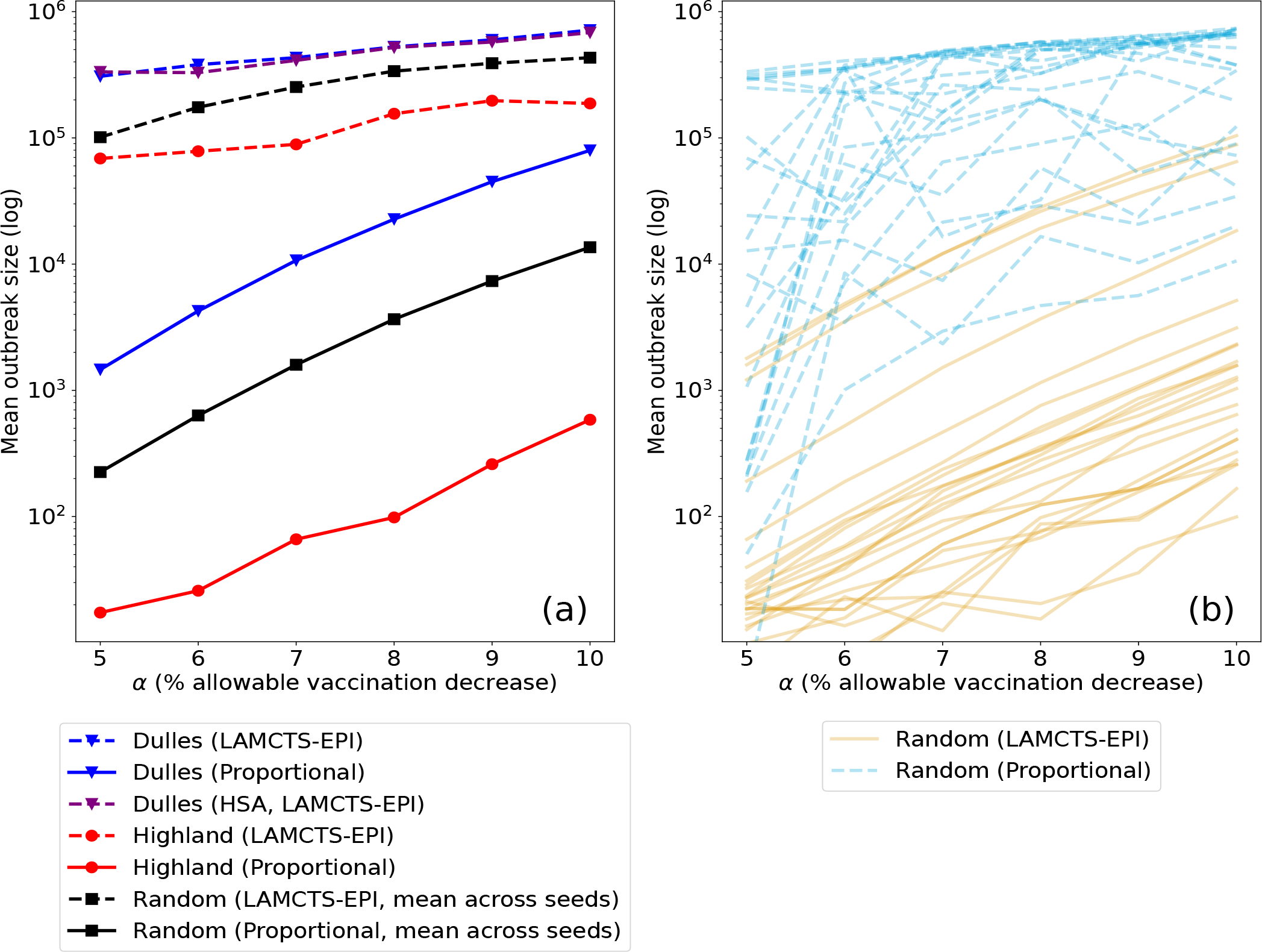
(a) Mean outbreak size (across 10,000 simulation replicates) versus *α*, the allowable statewide vaccination decrease for all conditions studied. The dotted and solid lines denote results obtained by the LAMCTS-EPI and Proportional scenarios, respectively. All scenarios result in increase in outbreak size with respect to *α*. Dulles has the largest outbreak size, Random is the second largest, and Highland scenarios are the smallest. (b) Mean outbreak size for a sample of runs from the Random scenario in the Proportional and LAMCTS-EPI scenarios.

**Figure 3:**
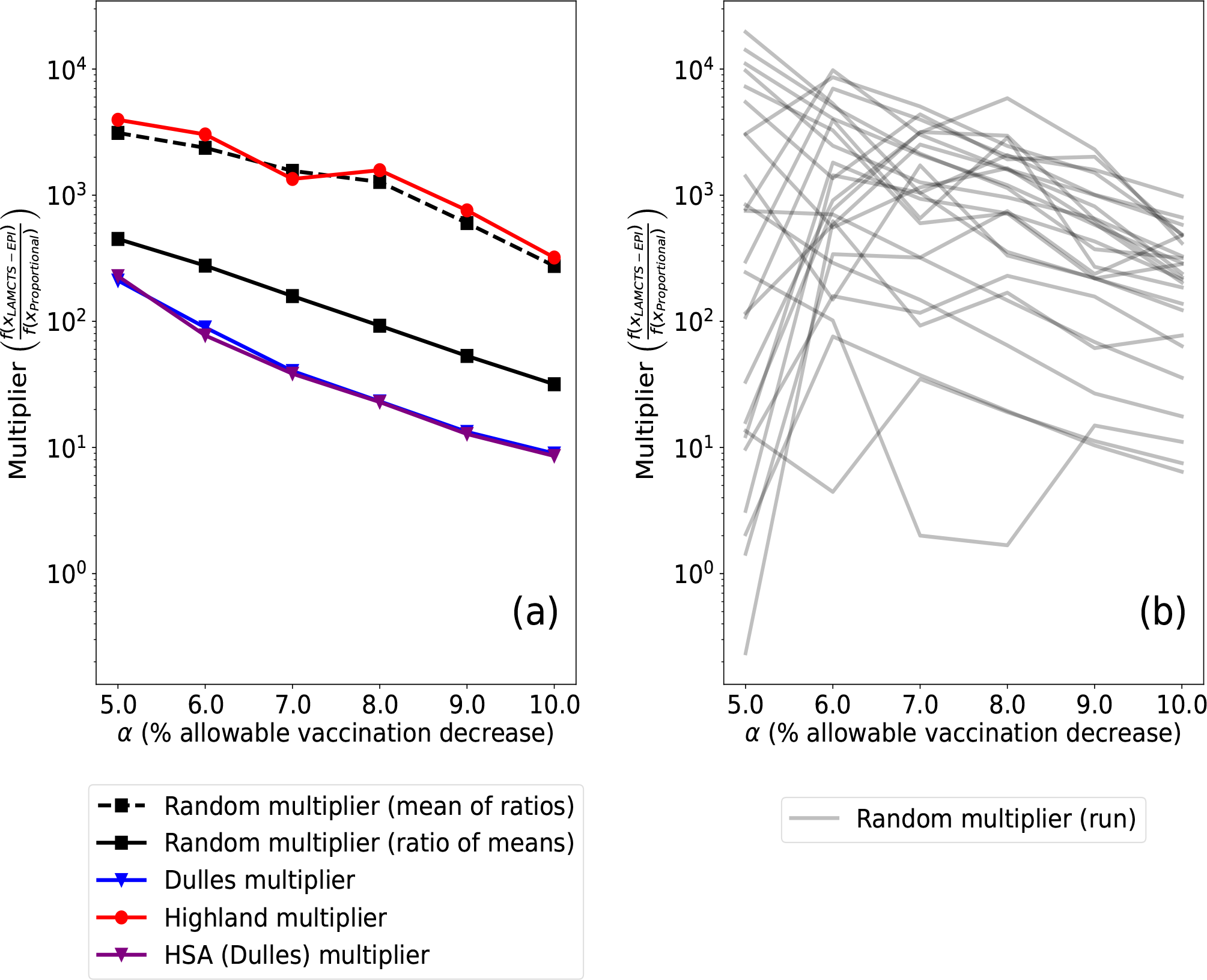
(a) Multipliers vs *α* for Dulles, Highland, HSA, and Random scenarios. There are two lines for Random, representing two different ways of summarizing the typical Random multiplier result. ‘Mean of ratios’ involves computing the multiplier for each run, then taking the mean of the computed multipliers. ‘Ratio of means’ involves computing the mean outbreak across all 25 seeds for the LAMCTS-EPI and Proportional scenarios, then taking the ratio. (b) Multipliers for each individual random run.

**Figure 4:**
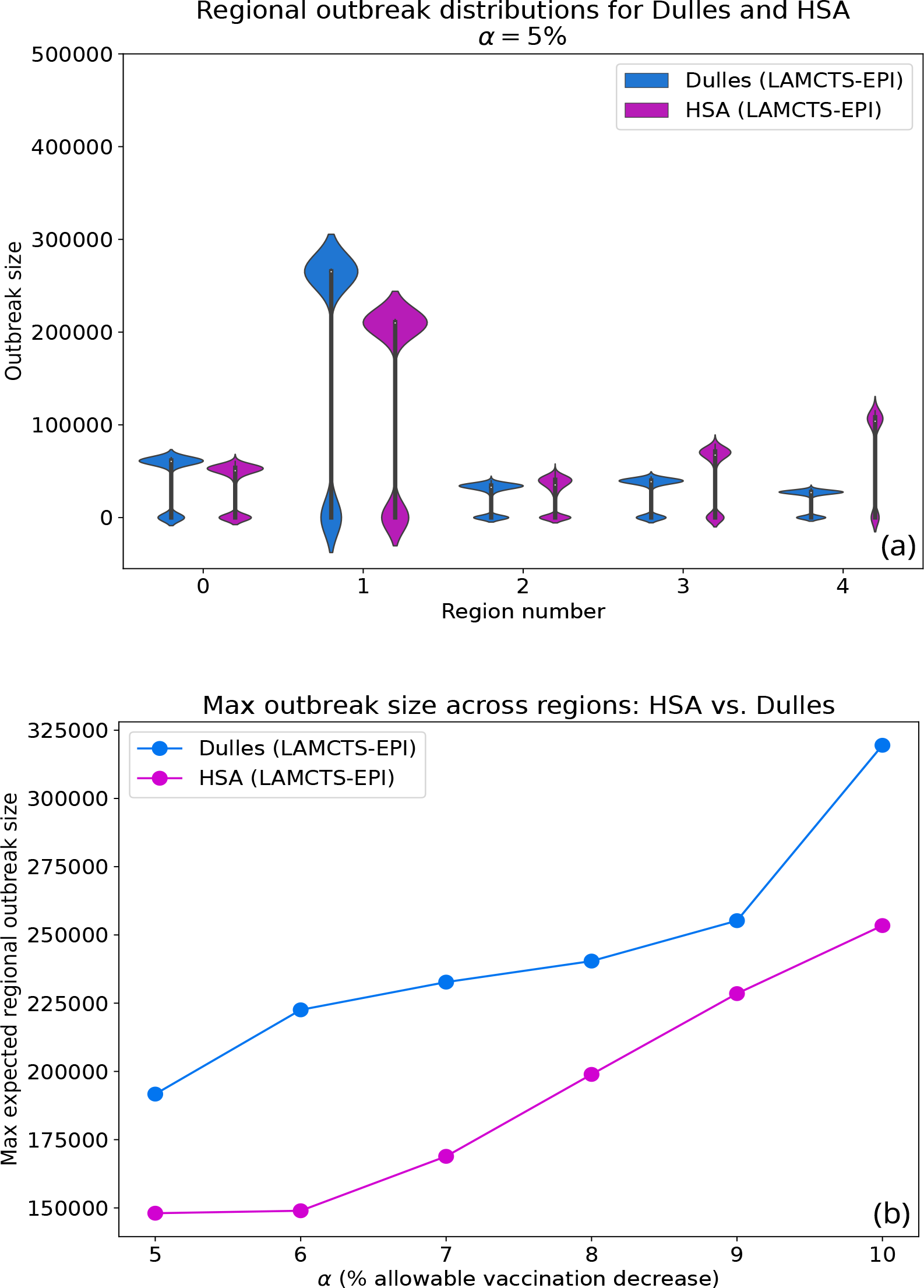
(a) shows the outbreak distribution for each HSA region in both the Dulles LAMCTS-EPI and HSA scenarios. We see that HSA has a smaller “large mode” than Dulles in region 1, but has outbreaks with similar or larger size than Dulles in other regions. (b) shows the maximum regional expected outbreak size. This is computed by calculating the expected outbreak size for each region, then taking the maximum expected outbreak size across the regions. For the *α* considered, the region with the highest outbreak size was usually region 1, which corresponds to the Northern Virginia region and contains the seeding location in this scenario. The HSA scenario has a smaller maximum regional expected outbreak size.

**Figure 5:**
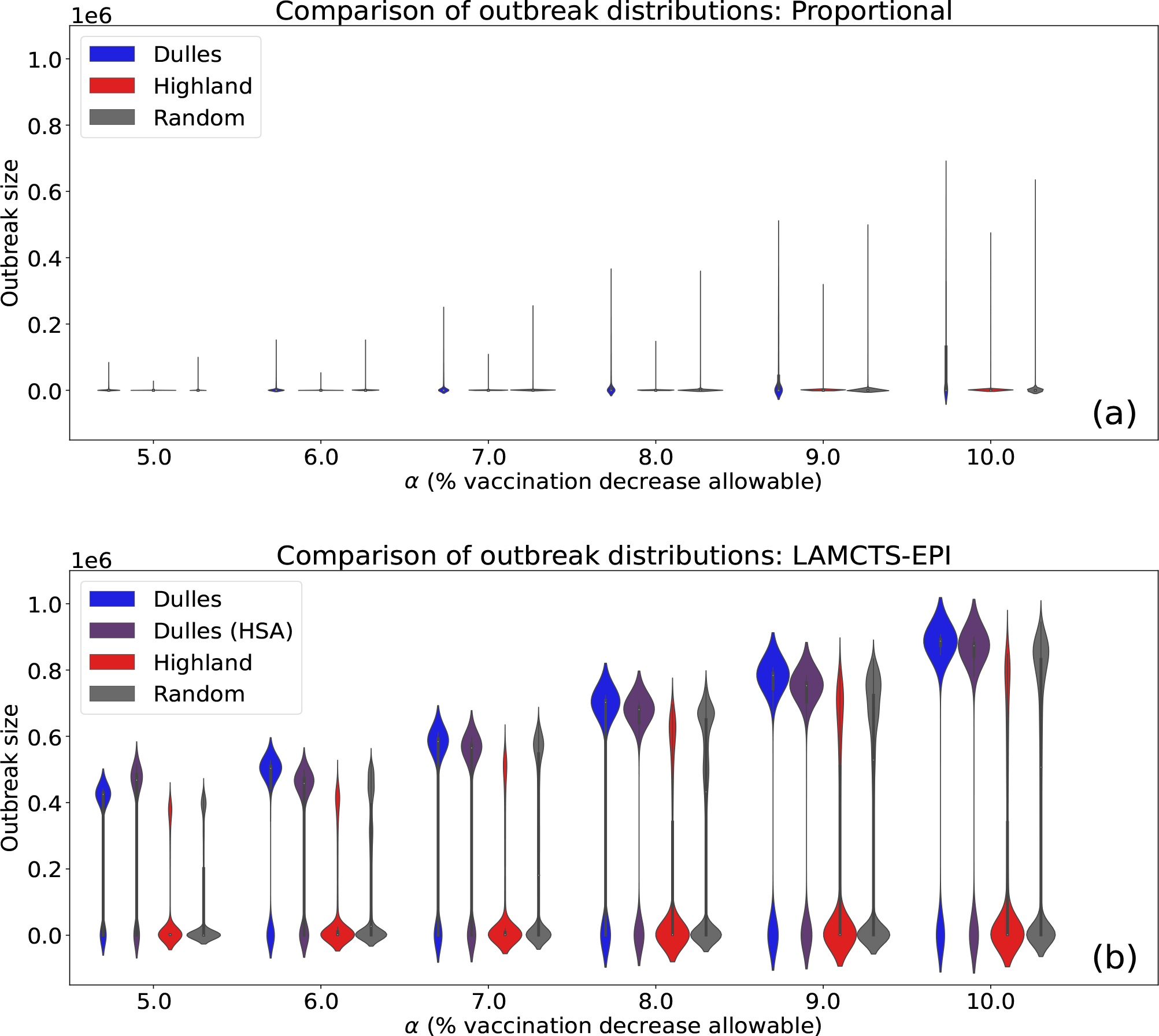
(a) Outbreak distributions (estimated from 10,000 simulation draws) of Proportional scenario versus *α*, the statewide percent allowable decline in vaccination. (b) Outbreak distributions (estimated from 10,000 simulation draws) of the LAMCTS-EPI scenarios vs. *α*, the statewide percent allowable decline in vaccination. Proportional distributions are long-tailed with a concentration around 0, indicating most simulations do not result in outbreaks. LAMCTS-EPI distributions are bimodal, with one mode around 0 and another mode at a large outbreak size. The mode corresponding to the large outbreak size appears to increase roughly linearly with respect to *α*. The large mode are fairly close to each other across all conditions.

**Figure 6:**
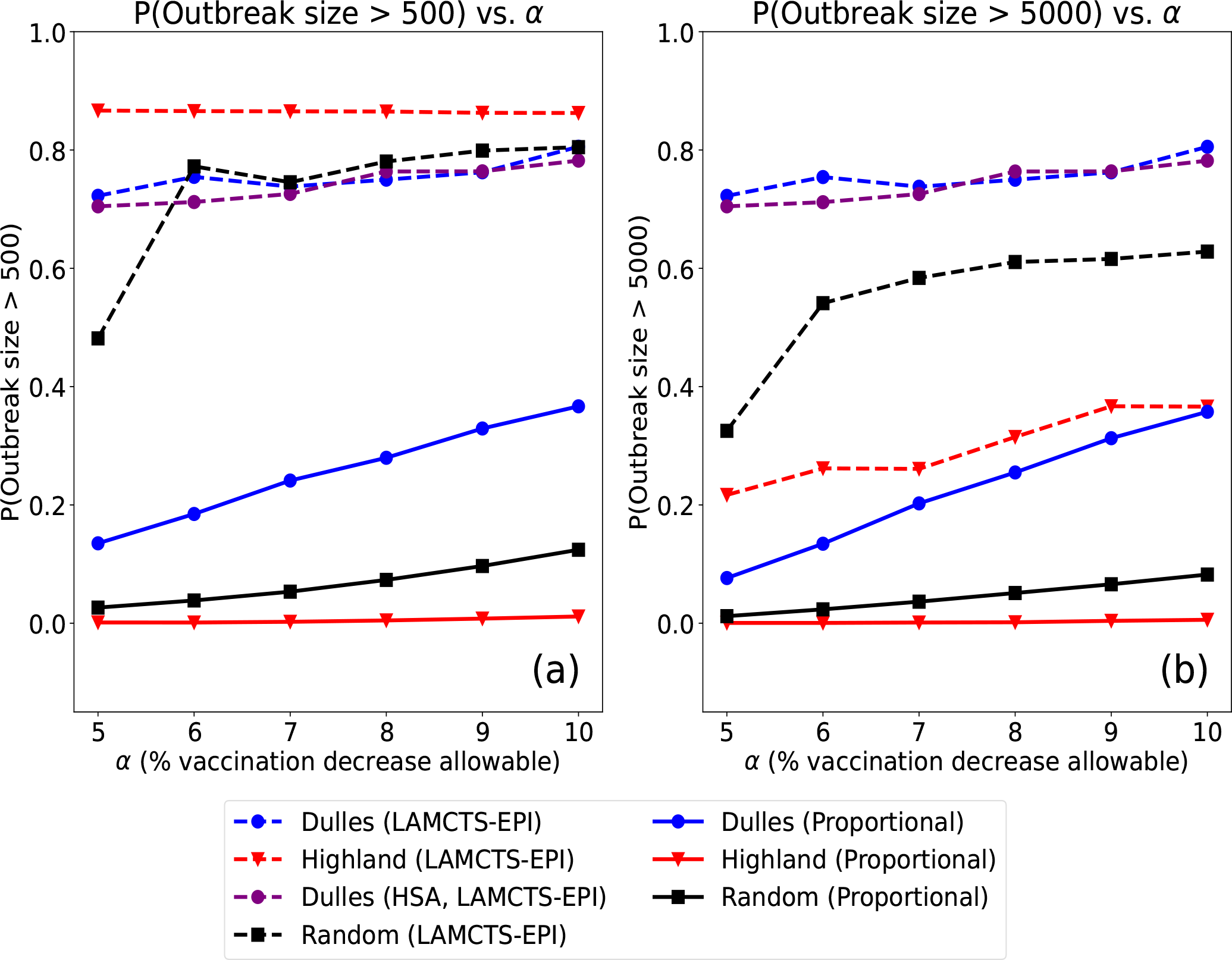
(a) Probability that total outbreak size exceeds 500 cases versus *α*. (b) Probability that total outbreak size exceeds 5000 cases versus *α*. As observed in Figure 2, probability increases w.r.t. *α* for both Proportional and LAMCTS-EPI scenarios. In the Proportional scenarios, Dulles has the largest outbreak probability, followed by Random and Highland. On the other hand for the LAMCTS-EPI scenarios: for low outbreak threshold (500), Highland has the largest probility, while Dulles and Random have similar probilities (aside from *α* = 5%. This flips for larger threshold of 5000: Dulles has the largest outbreak probability, followed by Random and then Highland. The Dulles probability nearly does not change, but Highland and Random drop considerably.

#### 3.1.1 Very significant risk of outbreaks due to decline in vaccination: effect of changing α

All lines on Figure 2 increase with respect to *α*. This is to be expected. An increase in *α*, or the decline in vaccination, should lead to more cases overall. However, the rates of growth in the Proportional versus LAMCTS-EPI scenarios is significantly different. All the Proportional scenarios have exponential growth with respect to *α*, with similar rates, increasing roughly one to one and a half orders of magnitude from *α*=5% to *α*=10%.

On the other hand, the LAMCTS-EPI scenarios show a much smaller increase with respect to *α*. However, they are more than two orders of magnitude higher than the Proportional solutions, especially for *α* = 5%. The gap between the LAMCTS-EPI and Proportional solutions reduces as *α* increases, and the gap at *α* = 10 is about an order of magnitude less than that at *α* = 5. In order to understand this behavior, we consider the multipliers, which is the relative gap between the LAMCTS-EPI and Proportional solutions, as defined in Section 2.4. In Figure 3, we observe the multipliers are all very high even at *α* = 5%, and decrease with *α. This implies that even at α* = 5, *the worst case outbreak risk is already very high*, irrespective of where the outbreak starts. This can be a very significant issue since immunization decline during the COVID period is reported to be at least at that level.

#### 3.1.2 Outbreak distributions

Figure 5 provides more insight into the exact nature of the outbreak distributions across these different scenarios. We see that the Proportional distributions are long-tailed with a sharp peak at 0, indicating most simulations do not result in large outbreaks. The tail grows longer as *α* increases, indicating that larger outbreaks do occur as *α* increases. However, they appear to still be relatively rare.

On the other hand, the LAMCTS-EPI distributions are bimodal with one mode at 0 and another mode with a large outbreak size. The Dulles scenarios have more mass near the mode associated with the large outbreak size and relatively less mass near 0. This is flipped for the Highland and Random scenarios, which have more mass near 0 compared to the mode at large outbreaks. For the LAMCTS-EPI scenarios, we see that the larger mode increases roughly linearly with respect to *α*. Overall, the modes for all conditions considered are quite close to each other.

Figure 4(a) shows that the regional outbreak distributions for the HSA and Dulles LAMCTS-EPI scenarios have a similar shape to the overall outbreak distributions. with each region having bimodal outbreak distributions. The maximum outbreak size for HSA is smaller compared to the maximum outbreak size of Dulles LAMCTS-EPI in region 1; but for all other regions, HSA either has comparable maximum outbreak size or larger. Figure 4(b) quantifies the effect of HSA on the maximum expected regional outbreak size. This is computed by first computing the mean outbreak size for each region and then taking the maximum of the regional means. The HSA maximum expected regional outbreak size is smaller than Dulles LAMCTS-EPI for all *α* considered.

#### 3.1.3 Outbreak probabilities

Figure 6 compares the probability that the outbreak size will exceed a given threshold with respect to *α*, for all scenarios. Two different outbreak thresholds are considered: 500 and 5000. All scenarios (Proportional and LAMCTS-EPI) have overall increasing probability of outbreak with respect to *α*, which makes sense considering Figures 2 and 5. For the Proportional outbreaks, the ordering is similar between the 500 and 5000 plot. Dulles Proportional has the greatest probability of exceeding the fixed threshold, followed by Random, then Highland. This ordering corresponds to the outbreak sizes in Figure 2. On the other hand, we see that the ordering changes when the threshold is raised for the LAMCTS-EPI results. At the lower threshold of 500, Highland LAMCTS-EPI has the highest probability of exceeding the threshold, whereas Random and Dulles have similar probabilities of exceeding the threshold. At the higher threshold, Dulles has the highest probability; and Random and Dulles have significantly reduced probabilities, with the ordering once again matching the log-outbreak plot. Notice that the line for Dulles is nearly identical between the two plots. One way of explaining this trend is to notice the amount of probability mass in between the modes of the distribution: Highland and Random have more mass in the middle and Dulles has nearly none, see Appendix **??**.

#### 3.1.4 Effect of importation

Figures 2 and 5 show that the seeding of the outbreak has a very significant impact on the outbreak risk, and it grows with *α*. The Dulles Proportional scenarios lead to a much higher expected outbreak size than the other scenarios, which is not surprising, given that zip code 20166 (where Dulles airport is located) is very urban, and has high population density and mixing. Similarly, the Highland scenarios have the lowest outbreak size, which is consistent with the low density in that county. On the other hand, we observe the multiplier is highest for Highland, starting at nearly 4 orders of magnitude at *α* = 5%, and reducing to roughly 3 orders of magnitude at *α* = 10%. For Dulles, the multiplier reduces from about 2.5 orders of magnitude to roughly 1 order of magnitude, as *α* varies from 5% to 10%. We present more details about the multipliers in Appendix **??**.

However, it is surprising that all the LAMCTS-EPI scenarios (including Highland) have pretty high outbreak sizes, and the gap between the scenarios is less than an order of magnitude, significantly less than the gaps in the Proportional scenarios. The distributions show a similar behavior in Figure 5.

#### 3.1.5 High variability in Random scenarios

Figure 2(a) shows the mean outbreak size (averaged across 10,000 simulations) for the Random Proportional and LAMCTS-EPI scenarios (along with the other scenarios), while Figure 2(b) shows a sample of individual runs, corresponding to random initializations.

We observe that there is a considerable amount of variance across the individual Random runs, both for Proportional and LAMCTS-EPI scenarios. However, the variance across the LAMCTS-EPI scenarios seems greater. The Proportional scenario lines concentrate in two different regions of the plot. The majority of the lines concentrate around the solid red Highland Proportional line, and there are a few that concentrate around the solid blue Dulles Proportional line (in Figure 2(a)). Thus, the pointwise mean of these lines lies between the Highland and Dulles lines. On the other hand, the Random LAMCTS-EPI scenario lines concentrate near the top of the plot, with considerable spread that stretches down into the middle of the plot. The multipliers for Random scenarios also have high variablity, as shown in Appendix **??**.

The higher variance is likely due to the additional sources of stochasticity present in the LAMCTS-EPI scenarios. The Proportional scenarios have two sources of randomness: (1) the stochastic nature of the tSIR simulation (which arises for all other scenarios), (2) the random selection of the seeding location (which is missing in the Dulles and Highland scenarios). In addition to these sources, the Random LAMCTS-EPI scenarios have an additional source: (3) the modified LA-MCTS optimizer.

Figure 3 considers two separate ways of computing the multiplier for the random scenario. One option is to take a “mean of ratios” -i.e, for the LAMCTS-EPI and Proportional run for seed *i*, first compute the ratio and then take the mean: 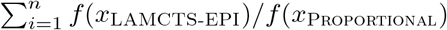. Alternately, one can also consider a “ratio of means” where first the mean LAMCTS-EPI and Proportional is averaged across seeds and then the ratio is taken: 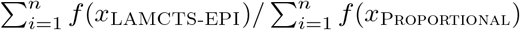. These quantities turn out to have a notable difference. The “ratio of means’ lies between the Highland and Dulles multiplier lines, whereas the “mean of ratios’ turns out to correspond closely to the Highland line.

This can be interpreted as follows: the “mean of ratios” gives evidence about the typical performance of the optimizer. In other words, if a random zipcode is selected as a seeding location and we compare its Proportional outbreak to its LAMCTS-EPI outbreak, it is likely that its multipliers will be similar to Highland’s. This may be a consequence of the fact the majority of zipcodes in VA are in more rural areas. On the other hand, the “mean of ratios” gives evidence about the gap between the average baseline size versus the average worst-case size. Considering all random Proportional runs as one “pool” of runs and all random LAMCTS-EPI runs as another “pool”, the ‘mean of ratios’ then gives information about the relative size of the means of these “pools.” In a sense, this is a restatement of what we see in Figure 2 with respect to how the Random means sit in between Highland and Dulles.

#### 3.1.6 The MaxRiskConstrained problem: worst case outbreak risk under surveillance in different regions

Recall that the MaxRiskConstrained problem specifies the decline rate *α*_*i*_ for each surveillance region *s*_*i*_, which is a HSA region in our study. Figures 2 and 5(b) show the outbreak size for Dulles importation when *α*_*i*_ = 5% for all surveillance regions *s*_*i*_. We note that there is very little difference between this and the Dulles LAMCTS-EPI scenario. The actual distributions of outbreaks are also quite similar, as shown in Figure 5(b). This is also consistent with the multipliers for the HSA and Dulles scenarios being very close, as shown in Appendix **??**.

### 3.2 Characterization of the regions with high decline in LAMCTS-EPI solutions

We analyze the structure of the solutions produced by LAMCTS-EPI. Specifically, we explore which counties have high declines (referred to as critical regions) in these solutions, and how they are related.

#### 3.2.1 Spatial distribution

Figure 7 shows heatmaps of county level declines in vaccination levels in the LAMCTS-EPI solutions for the Dulles and Highland scenarios. We observe that the solutions vary significantly with the importation, and there are high declines generally close to where importation occurred. In the Dulles scenarios (Figure 7(a) and (b)), the declines are higher in the regions close to Loudoun county (20166), including Fairfax and Arlington. In the Highland scenarios (Figure 7(c) and (d)), the declines are positive in Highland county, but are higher a bit further, around Albemarle county. Further, in these solutions, there is not much decline in Loudoun county, and regions around it. In both these scenarios, we do notice some high declines spread across the state, quite far from the importation. This implies the mixing across the state plays a significant role.

**Figure 7:**
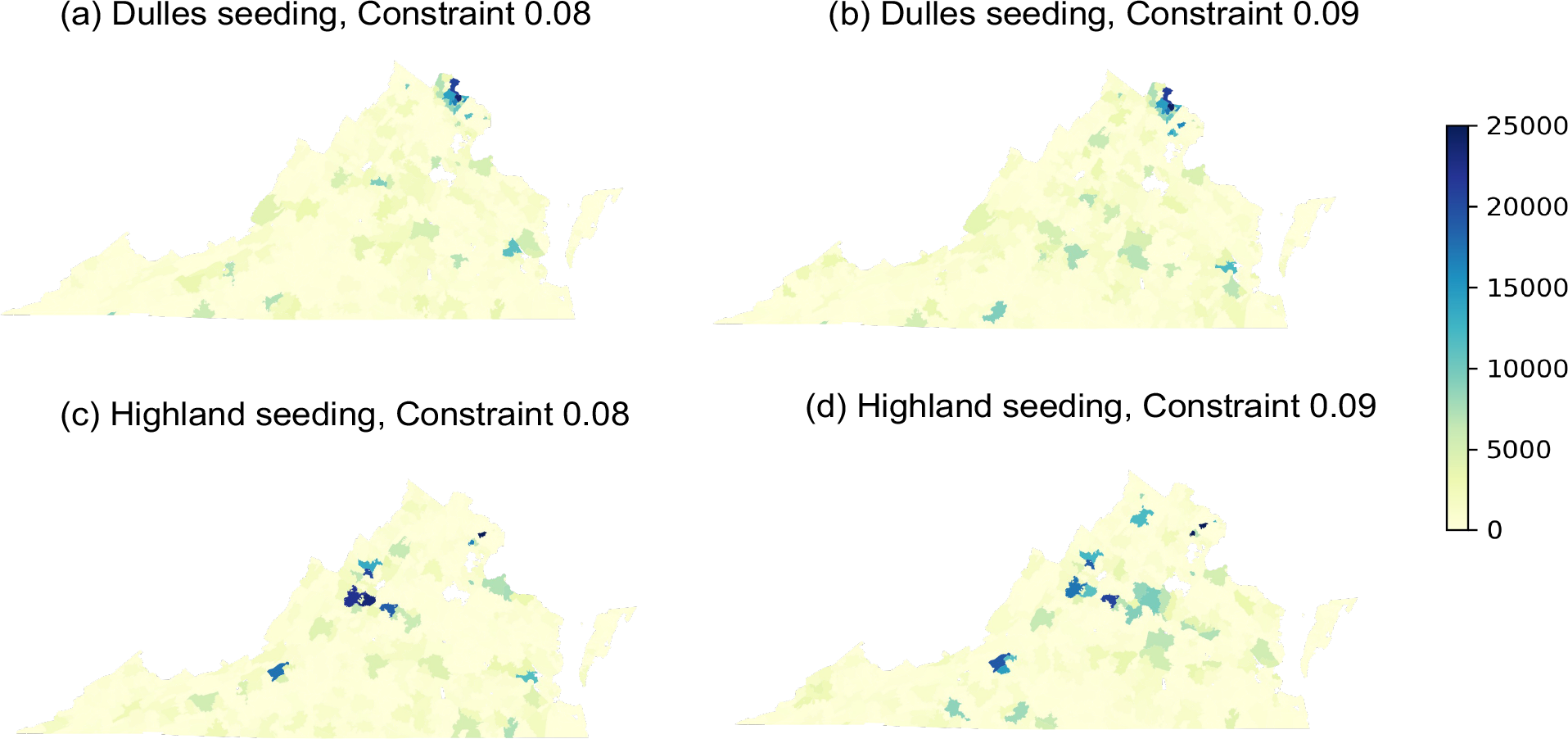
Spatial distribution of the unvaccinated population in the LAMCTS-EPI solutions for the Dulles and Highland scenarios for *α* equal to 8% and 9%.

We find that the distribution of county level decline in vaccination changes when *α* changes. For instance, in Figure 7, when *α* changes from 8% to 9%, new regions appear, and some of the earlier regions are dropped. We note that many of the counties with high declines continue when *α* is increased. For Dulles seeding, 54.7% counties show higher decline from 8% to 9% and the rest of the counties show lower decline. For Highland seeding, 53.1% counties show higher decline from 8% to 9% (Figure 8).

**Figure 8:**
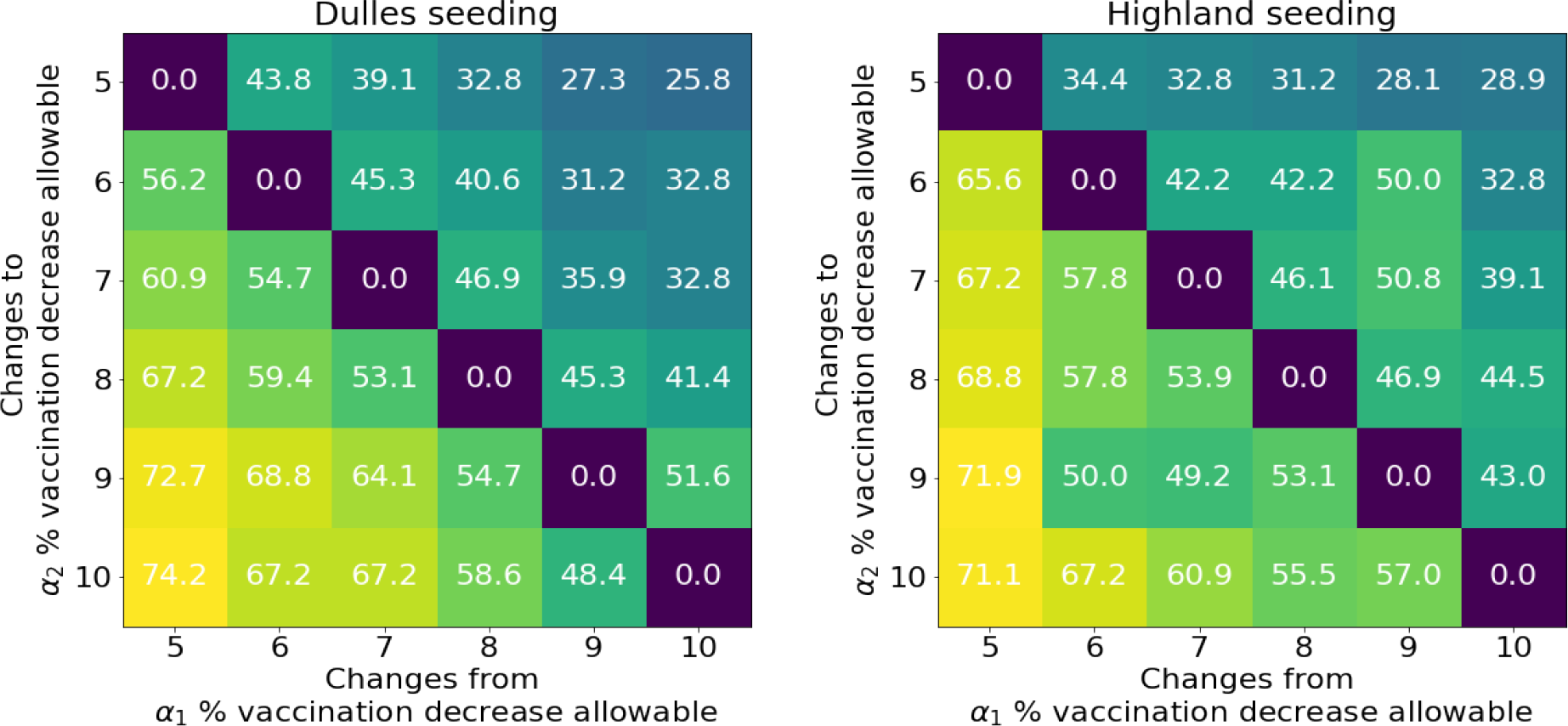
Percentage of counties where the unvaccinated population increases as *α* transitions from *α*_1_ to *α*_2_.

#### 3.2.2 Clustering indices

We compute the Isolation index and Moran’s-I index for the LAMCTS-EPI solutions for all importations; for Random, we average these indices over the random seedings. If there is not much separation between regions with high decline, the isolation index would be close to the average decline (across counties), which equals *α*. We find that the isolation index is much higher for each importation scenario in the optimizer output, as shown in Figure 9(left), which indicates that the zip code level declines are concentrated in a few geographic locations. Among the three seeding scenarios, Highland has much higher isolation index for all *α*.

**Figure 9:**
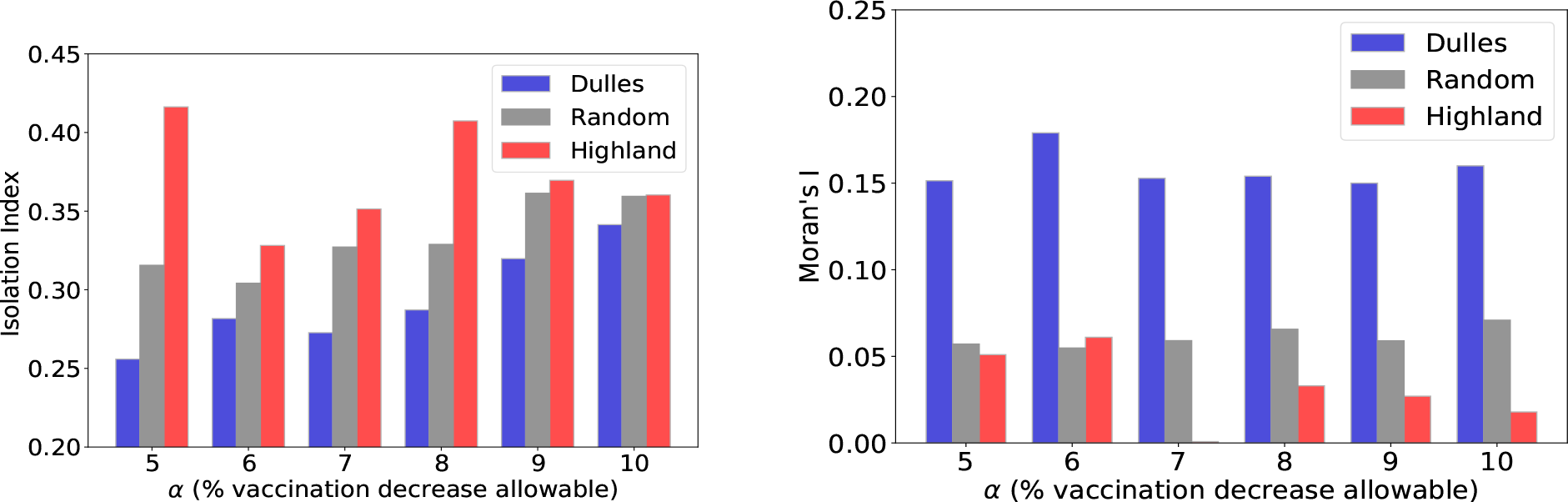
Isolation Index and Moran’s I for different seeding scenarios.

On the other hand, we find that Moran’s I is quite low for all the scenarios, as shown in Figure 9(right), which indicates no association in spatial distribution of the zip code level drops in vaccination for Random and Highland importation. However, Dulles importation indicates high-high zip code level distribution of the unvaccinated population.

Figure 9(left) and Figure 6 show that higher isolation index has associativity with the potential outbreak size. Therefore, this metric can be useful to understand the outbreak vulnerability of a location.

#### 3.2.3 Characterizing the LAMCTS-EPI solutions

We examine if the county level decline in vaccinated individuals in the LAMCTS-EPI solutions has any associations with different county level features, such as: the county’s population, immunization rate, vaccinated population, and other characteristics derived from Census, such as, gender, race. Our regression analysis finds all predictors to be statistically insignificant in explaining the decline in the number of vaccinated individuals in the county (Table **??** in Appendix).

## 4 Discussion

### Effectiveness of our optimization techniques

The MaxRisk and MaxRiskConstrained problems are computationally very challenging when the epidemic model is complex. We find that our LAMCTS-EPI algorithm, and the Bayesian optimization techniques are quite effective in finding good solutions to these problems; further, the solutions are significantly different, compared to other baselines. In particular, the actual patterns of decline in immunization, mixing patterns and importation have a very significant impact on the outbreak risk. While the role of patterns of decline was noted in [28], solving a formulation such as MaxRisk, and an optimization framework such as LAMCTS-EPI, are necessary for understanding the worst case outbreak risk in more complex disease models. Our optimization framework is very general, and can be easily adapted to other public health problems.

### Very significant risk of outbreaks due to decline in vaccination rates

Our main observation is that decline in vaccination rates poses very high risk of outbreaks. Even at the lowest level of *α* = 5%, we find the expected outbreak size is in the thousands in the Dulles Proportional scenario (though it is much smaller for Random and Highland Proportional scenarios). The outbreak size grows exponentially with *α* in the Proportional scenarios. However, the worst case outbreak size in the LAMCTS-EPI scenarios is multiple orders of magnitude higher than the Proportional scenarios, but have limited variation with *α*. This means that even for *α* = 5%, the worst case outbreak risk is already quite high, and could be a significant concern for public health agencies. Our results also show that there are different “motifs of vaccination drop” (following the concepts introduced in [28]), which depend on the connectivity structure in a complex manner. At a qualitative level, this is consistent with the results of [11], who show that spatial heterogeneity in immunization rates has a significant impact on the outbreak size. In particular, they find that spatial heterogeneity in immunization rates can increase the epidemic size when the population has a sufficiently high rate of vaccine coverage. This is consistent with the outbreak we observe around Loudoun county, though we don’t see very specific patterns in the worst case solutions.

### Significance of importation location

We find importation leads to significant differences with respect to all the three metrics we consider, namely, expected outbreak size, multiplier, and probability of outbreak. Our results confirm our basic intuition about importation: seeding in Loudoun county (Dulles scenarios) poses a much higher risk as it causes ‘larger than average’ outbreaks both in terms of frequency and size. Seeding in Highland county causes a ‘smaller than average’ outbreak. In terms of the absolute outbreak size, the Dulles scenario has the worst outbreak size.

Figure 3 shows that the Dulles scenario has a much smaller multiplier compared to Highland scenario. A possible explanation for a lower multiplier in Dulles is that the pool of susceptible is exhausted even without optimization. Since Loudoun county, containing Dulles, is considerably more dense compared to the rest of VA, a 5% reduction in vaccination rate results in a fairly large number of susceptible individuals in the area. This is enough “mass” to cause fairly large outbreaks in the baseline scenario itself, and there are not that many more susceptibles available to infect in the worst case scenario. This is in contrast to Highland, which does not have the same mass in the baseline and hence optimization allows many more susceptibles to be infected.

We also find significant differences in the probability of an outbreak. For a lower outbreak size threshold of 500 infections, Highland has the highest probability, but for a higher threshold of 5000, the probability is much higher for Dulles. Possible reasons are: Loudoun county is a part of a large metropolitan area with high population density so a small spark can spread quickly and cause a big outbreak. On the other hand, Highland, although has a lower vaccination rate, which makes initial infections more likely, but given the population size and density, the probability of it spreading is quite low.

### Outbreak risk under surveillance in different regions

We find that the overall outbreak sizes to the MaxRiskConstrained and MaxRisk problems for the Dulles scenario do not differ much for *α* = 5%. This means that even if the decline is known to be 5% in each HSA region, the outbreak size is basically the same as if the decline is 5% statewide. One way to understand this is that since the optimizer can no longer attain large outbreak sizes by concentrating declines in one highly populous region near the seeding location, it “compensates” by shifting the decline to other metro areas within separate health regions. For example, we find that the Virginia Beach metro area, located within southeast Virginia, experiences a larger decline in the HSA scenario. Appendix **??** shows heatmaps of the solutions obtained by LAMCTS-EPI for *α* = 5 in the HSA and Dulles LAMCTS-EPI scenarios.

While the addition of constraints failed to affect the statewide outbreak size, we do find that it can reduce the concentration of risk within a single region. As the optimizer shifts the risk from one region to another, the maximal outbreak size that any particular region might experience is decreased relative to the unconstrained case. On the other hand, this is associated with a marginal increase in risk in other regions. An interesting point is that the outbreak size in the Dulles scenario for MaxRisk does not vary much with *α*. Therefore, it is possible that even for a slightly smaller *α*, most susceptibles around the importation region are exhausted. We suspect there would be more differences between the MaxRisk and MaxRiskConstrained solutions for non-uniform *α*_*i*_ values, and future work is needed to understand the impact of non-uniformity.

### Characterization of declines in different regions

Our results show the absence of any significant associations between the county level decline in the vaccinated individuals in the LAMCTS-EPI solution and various county level characteristics (such as the size of the population and different sub-populations). This needs further investigation, but there could be multiple reasons. It is possible that mixing across counties based on the gravity model is playing a bigger role than other demographic features. It may also be the case there are many such bad solutions which have high outbreak size; we only consider the worst solution computed by LAMCTS-EPI. This may also be connected with the lack of clustering with respect to Moran’s-I. We believe this is an important topic for future study.

### Implications for healthcare policy and planning

For planning purposes, the metrics we study, along with the demographic and geographical factors, can provide valuable information to the public health authorities. Dulles, situated in suburban/urban region in Northern VA, a relatively wealthy area of VA, is likely to have more health resources. Whereas, Highland, a rural area, is unlikely to be resource rich. Also, our model assumes no interventions. So, if an outbreak were to occur in Dulles vs. Highland - it may be worse for an average resident of Highland than it would be for an average resident of Dulles area because the health agencies in Dulles may be better equipped to intervene compared to Highland. This points to the fact that describing risk of a cluster is not entirely as simple as “maximal expected outbreak risk”. Kundrick et al. [26] explored the usage of different criteria (vaccination coverage, epidemic risk, susceptible birth cohort) for vaccine prioritization in Malawi given different goals. They pointed out that the measles outbreak in Malawi in 2010, began and spread fastest in regions around Blantyre, where vaccination coverage was estimated to be high. Similar multi-criteria objectives and priorities need to be considered when applying our results in a public health planning context.

### Limitations and future work

The specific results in this study are for the tSIR model [45]. This has been used extensively, especially for measles modeling. More generally, *β* is assumed to be the same for all patches at all times, but this may not necessarily be the case in reality. For example, Xia et al. [45] explored the possibility of time-varying *β* dependent on upon school schedules, when sustained measles transmission was most likely. Bjørnstad et al. [6] also found that the *β* parameter was dependent upon city size when the tSIR model was used to infer parameters from historical incidence data. Other modeling approaches have also been considered for measles, including agent based models, e.g., [43]. The specific results and outbreak estimates might change with these disease model. However, we expect the general conclusions will still hold. As there has been no outbreak in Virginia, our results are also conditional on the calibration using a scaling of the New York outbreak. Such risk estimation analyses for other regions might have similar issues, since there have not been any large outbreaks in most regions.

Another important modeling assumption in our study is that no interventions are implemented. This is clearly unrealistic, since infected individuals would be isolated. Our results are likely to give conservative upper bounds, as in [28], who also consider a simple disease model without interventions. We expect the actual outbreak size and other metrics would be lower if interventions are implemented, but many of the results are likely to be qualitatively similar.

We note that in our results, LAMCTS-EPI is setup for the expected outbreak size objective. The results for other objectives, such as the outbreak probability could be quite different. In addition, model calibration could give a distribution over models, and solutions whose objective value is high over the distribution might have a different structure. Another consideration for exploring the solution space is to consider sets of solutions rather than searching for a single solution. Recent work by Maus et al. [29] explored the possibility of finding many solutions whose objective value exceed a predefined threshold. This approach might enable the discovery of solutions with a variety of different structures.

## 5 Conclusions

The MaxRisk and MaxRiskConstrained problems formalize the outbreak risk due to decline in vaccination rate, based on available survey information, which can be at different levels of resolution. Both problems are motivated by the work of [28], who show that how the immunization rates are spread within a region (which they refer to as vaccination motifs) could have an impact on the outbreak risk. Both these problems are very challenging due to the large decision space, and the non-convex objective. Our algorithm, LAMCTS-EPI, builds on powerful Bayesian optimization techniques, and finds good solutions to these problems.

Our algorithm enables finding solutions for state level models. Our results on a model for Virginia show that the worst case outbreak risk, computed by LAMCTS-EPI, even for a 5% drop in immunization rates, is very high. This would not be identified by assuming proportional drop in immunization across counties, which is consistent with the observation of [28], who showed that vaccination motifs have a significant impact on the outbreak size. Our results also show that the actual mixing patterns and importation have a very significant impact on the outbreak risk. The LAMCTS-EPI solutions have high declines in counties close to the importation regions, but not very consistently. We also do not see any significant associations between the decline in the optimal solutions and many county level features. Since surveillance data on vaccination rates is generally available only at a coarse resolution, our methods provide a systematic way of understanding the future risk of outbreaks of vaccine preventable diseases.

## Data Availability

All data are publicly available

## 6 Author contributions

**Nicholas Wu**: Investigation, Methodology, Software, Writing - Original Draft, Writing - Review and Editing; **Sifat Afroj Moon**: Conceptualization, Investigation, Methodology, Software, Writing - Original Draft, Writing - Review and Editing; **Ami Falk**: Software, Methodology; **Achla Marathe**: Conceptualization, Writing - Review and Editing; **Anil Vullikanti**: Conceptualization, Writing - Original Draft, Writing - Review and Editing.

## 7 Acknowledgements

## Footnotes

1 https://apps.vdh.virginia.gov/HealthStats/documents/2010/pdfs/HDMap.pdf

